# Regional atrophy and cognitive decline depend on definition of subjective cognitive decline

**DOI:** 10.1101/2021.04.21.21255752

**Authors:** Cassandra Morrison, Mahsa Dadar, Neda Shafiee, Sylvia Villeneuve, D. Louis Collins, for Alzheimer’s Disease Neuroimaging Initiative

## Abstract

**Background:** People with subjective cognitive decline (SCD) may be at increased risk for Alzheimer’s disease (AD). However, not all studies have observed this increased risk. Inconsistencies may be related to different methods used to define SCD. The current project examined whether four methods of defining SCD (applied to the same sample) results in different patterns of atrophy and future cognitive decline between cognitively normal older adults with (SCD+) and without SCD (SCD-).

**Methods:** MRI scans and questionnaire data for 273 cognitively normal older adults from Alzheimer’s Disease Neuroimaging Initiative were examined. To operationalize SCD we used four common methods: Cognitive Change Index (CCI), Everyday Cognition Scale (ECog), ECog + Worry, and Worry only. A previously validated MRI analysis method (SNIPE) was used to measure hippocampal *volume* and *grading*. Deformation-based morphometry was performed to examine volume at regions known to be vulnerable to AD. Logistic regressions were completed to determine whether diagnostic method was associated with volume differences between SCD- and SCD+. Linear mixed effects models were performed to examine the relationship between the definitions of SCD and future cognitive decline.

**Results:** Results varied between the four methods of defining SCD. Left hippocampal grading was lower in SCD+ than SCD-when using the CCI (*p*=.041) and Worry (*p*=.021) definitions. The right (*p=*.008) and left (*p=*.003) superior temporal regions were smaller in SCD+ than SCD-, but only with the ECog. SCD+ was associated with greater future cognitive decline measured by Alzheimer’s Disease Assessment Scale, but only with the CCI definition. In contrast, only the ECog definition of SCD was associated with future decline on the Montreal Cognitive Assessment.

**Conclusion:** The current findings suggest that the various methods used to differentiate between SCD- and SCD+ influence whether volume differences and findings of cognitive decline are observed between groups in this retrospective analysis.

## Introduction

In 2020, there were approximately 50 million people worldwide living with dementia. Alzheimer’s disease (AD) accounts for around 50-75% of these cases (1). People with AD experience progressive declines in cognitive functioning, including memory and thinking, with symptoms interfering with daily functioning (2). These symptoms may occur because of brain pathology, including excessive amyloid and tau build-up and neurodegeneration in regions associated with cognitive deficits (3). Recent work suggests that this neuropathology may be present for many years before the onset of behavioral symptoms that interfere with daily activities and cognitive functioning (4,5). Much of the current AD research has been devoted to finding an early biomarker that can identify individuals with preclinical AD.

People with preclinical AD are cognitively normal but display AD pathology (5). Individuals meeting the criteria for preclinical AD may also report subjective cognitive decline (SCD), i.e., perceived deficits in memory and/or cognitive functioning in the absence of objective cognitive decline (6). Reports of SCD may occur up to 15 years before the onset of AD symptoms (7) and increases the likelihood of progression to clinically probable AD by up to five times (8). SCD may be one of the earliest clinical manifestations of AD (6) with AD pathology including amyloidosis and neurodegeneration being observed in people with SCD (see 7,8 for review).

Much research has shown that people with SCD show increased neurodegenerative pathology compared to cognitively normal older adults without SCD. Nevertheless, results suggesting atrophy declines in people with SCD compared to those without SCD are inconsistent. For example, while Jessen et al. (10) observed atrophy in the bilateral entorhinal cortex and not the hippocampus, Striepens et al. (11) reported reduced volume in both the bilateral entorhinal cortex and bilateral hippocampus in people with SCD relative to people without SCD. Different methods used to classify SCD may underly the conflicting results. Jessen et al. recruited participants who sought medical help for their self-reported feeling of worsening memory with an onset in the last 5 years (10). On the other hand, Striepens et al. recruited participants who had both self and informant confirmation of memory impairment within the last 10 years (12). They may have observed reduced hippocampal in people with SCD because of including informant confirmation. Informant confirmation is a feature of SCD *plus* (people with SCD who have features that make them more likely to progress to AD) and might be a better predictor of objective performance as disease severity progresses (8,13). Therefore, the participants included in the Streipens et al. study may be closer to clinical decline than those recruited in the Jessen et al. study.

Inconsistencies in defining SCD lead to the development of the Subjective Cognitive Decline Initiative (SCD-I; 6). This group developed a broad definition of pre-MCI SCD for research, which includes self-experienced persistent decline in cognition compared to a previously normal status and normal performance on cognitive tests. MCI/AD, psychiatric conditions, neurological diseases, medical disorders, medications, or substance abuse cannot explain these self-perceived declines. Some other important features that may improve SCD identification include the study setting, *Apolipoprotein E (APOE*) status, memory vs non-memory domain complaints, informant confirmation, and worry. The authors provide specific features that increase the likelihood of preclinical AD (SCD *plus*) including 1) SCD in memory (rather than other domains), 2) onset within the last 5 years, 3) age of onset ≥60 years, 4) worry/concern with SCD, and 5) feeling of worse performance than others of the same age. They also note that for SCD *plus*, informant confirmation of decline, *APOE ε4* positivity, and AD biomarker evidence is important.

Despite the SCD-I working groups’ recommendations (6), standards and features in defining SCD have not been universally implemented. One concern is the lack of standardization of the methodologies used to capture SCD. The review by Wang et al. (9) provides further insight into this problem. Some studies use specific questionnaires to define SCD such as the Cognitive Change Index (CCI; 10), Everyday Cognition Scale (ECog; 12), or Memory Assessment Clinics Questionnaire (MAC-Q; 13), others use one or two questions (e.g., do you have memory declines; are you worried about those declines), while some use memory clinic consultation to define people with SCD. Several potential problems exist with this lack of standardization. Few studies have examined the relationship between different SCD questionnaires. One study found a moderate correlation between the ECog and Blessed Memory scale (17). Similarly, only a moderate correlation between the MAC-Q and the Subjective Memory Complaints (SMC) scale has been observed (18). These findings suggest that different SCD questionnaires may not be measuring the same construct and may influence the heterogeneous results regarding the association between SCD and early AD-biomarkers.

The goal of the current study was to examine whether brain atrophy and future cognitive decline observed between cognitive normal older adults with and without SCD vary between four methods of differentiating SCD. We also completed another analysis, including APOE ε4 status and amyloid positivity as two factors in the model because they may be associated with increased risk for AD (see 7 for review). In this study, we examined the following regions of interest (ROIs): hippocampus, amygdala, lateral ventricles, and superior temporal regions. While there are many regions that have been shown to be associated with AD-pathology, several studies have observed that the hippocampus may be sensitive to these changes in SCD+ populations (see (9) for review). When predicting who will convert to AD in cognitively healthy older adults, hippocampal changes have shown a prediction accuracy of 72.5% up to 7 years in advance (19). The superior temporal regions and amygdala were chosen for analysis as previous research has also shown that these regions have high prediction accuracy when examining conversion from SCD to MCI (20). The lateral ventricles have also shown to be sensitive to detection of early changes in MCI (21).

## Methods

### Alzheimer’s Disease Neuroimaging Initiative

Data used in the preparation of this article were obtained from the Alzheimer’s Disease Neuroimaging Initiative (ADNI) database (adni.loni.usc.edu). The ADNI was launched in 2003 as a public-private partnership, led by Principal Investigator Michael W. Weiner, MD. The primary goal of ADNI has been to test whether serial magnetic resonance imaging (MRI), positron emission tomography (PET), other biological markers, and clinical and neuropsychological assessment can be combined to measure the progression of mild cognitive impairment (MCI) and early Alzheimer’s disease (AD). The participants of this study are from the ADNI-2 cohort, between 55 and 90 years old at the time of recruitment. The ADNI-2 cohort participants were used because it was the first cohort to introduce the CCI questionnaire and define participants with significant memory concerns. For consistency with current research standards, we use the definition of subjective cognitive decline. The study received ethical approval from the review boards of all participating institutions. Written informed consent was obtained from participants or their study partner before the testing procedures began.

### Participants

While there are 420 cognitively normal controls in the ADNI-2 cohort, participants were only included in this study if they had an MRI scan within 6 months of SCD questionnaire completion and fully completed the questionnaires accessing SCD. A total of 280 total participants from the ADNI-2 dataset met the requirements for this study. The participants were separated into either cognitively normal controls without subjective cognitive decline (negative for subjective cognitive decline, or SCD-) or cognitively normal controls with subjective cognitive decline (positive for subjective cognitive decline, or SCD+)^1^. Both groups had no objective evidence of cognitive impairment on cognitive tasks or the Clinical Dementia Rating and no signs of depression. Four separate definitions of SCD were used to differentiate the participants into either the SCD- or SCD+ group; therefore, the number of SCD- and SCD+ participants varied between each analysis as the criteria for SCD differed.Demographic information, by group, for each definition is presented in Table 1.

**Table 1:** Demographic and clinical characteristics for cognitively normal older adults with and without subjective cognitive decline

| Demographic information | Analysis 1- CCI |  | Analysis 2-ECog |  | Analysis 3- ECog + Worry |  | Analysis 4- Worry |  |
| --- | --- | --- | --- | --- | --- | --- | --- | --- |
|  | SCD-<br>n=176 | SCD+<br>n=97 | SCD-<br>n=130 | SCD+<br>n=143 | SCD-<br>n=177 | SCD+<br>n=96 | SCD-<br>n=149 | SCD+<br>n=124 |
| Age | 73.50 ± 6.27 | 72.44 ± 5.59 | 72.56 ± 6.21 | 73.63 ± 5.86 | 72.94 ± 6.17 | 73.43 ± 5.84 | 72.97 ± 6.17 | 73.29 ± 5.93 |
| Education | 16.55 ± 2.54 | 16.77 ± 2.59 | 16.58 ± 2.65 | 16.67 ± 2.48 | 16.63 ± 2.57 | 16.63 ± 2.56 | 16.64 ± 2.52 | 16.61 ± 2.62 |
| AV-45 | 1.11 ± 0.18 | 1.13 ± 0.19 | 1.10 ± 0.16 | 1.13 ± 0.19 | 1.11 ± 0.18 | 1.14 ± 0.19 | 1.10 ± 0.17 | 1.14 ± 0.19 |
| APOE ε4+ | 51 (29%) | 29 (30%) | 38 (29%) | 42 (29%) | 54 (30%) | 26 (27%) | 44 (30%) | 36 (30%) |
| Amyloid Positivity | 61 (35%) | 42 (43%) | 46 (35%) | 57 (40%) | 59 (33%) | 44 (46%) | 49 (33%) | 54(44%) |
| Male sex | 86(48%) | 61 (41%) | 64 (49%) | 64 (45%) | 88(50%) | 38 (40%) | 83(51%) | 50(40%) |
| ADAS-13 | 9.05 ± 4.44 | 8.79 ± 4.23 | 8.79 ± 4.23 | 9.19 ± 4.19 | 8.93 ± 4.57 | 8.84 ± 3.77 | 8.99 ± 4.77 | 8.78 ± 3.67 |
| MoCA | 25.75 ± 2.37 | 25.65 ± 2.58 | **26.12 ± 2.35** | **25.36 ± 2.47\*** | 25.83 ± 2.37 | 25.50 ± 2.58 | 25.82 ± 2.36 | 25.58 ± 2.56 |
| MMSE | 29.02 ± 1.23 | 29.01 ± 1.22 | 29.12 ± 1.13 | 28.93 ± 1.30 | 29.06 ± 1.20 | 28.95 ± 1.28 | 29.08 ± 1.17 | 28.95 ± 1.30 |

Values are expressed as mean and standard deviation. APOE ε4+, amyloid positivity, and male sex are represented as total number of sample and percentage of sample. CCI = Cognitive Change Index. ECog = Everyday Cognition Scale. SCD- = Cognitively normal older adults without subjective cognitive decline. SCD+ = cognitively normal older adults with subjective cognitive decline. ADAS-13 = Assessment Scale–Cognitive Subscale. MoCA = Montreal Cognitive Assessment. MMSE = Mini Mental Status Examination.

For Analysis One, participants were identified as SCD+ if they had self-reported significant memory concern as quantified by a score of ≥16 on the first 12 items (representing memory changes) on the CCI. For Analysis Two, the ECog (14) was used to differentiate between SCD- and SCD+. If a participant endorsed any item on the ECog with a score ≥ 3 (signifying consistent SCD) they were assigned to the SCD+ group. The SCD-group consisted of those who selected either better, no change, or questionable/occasionally worse. The third analysis separated groups base on the ECog scale and Worry. The SCD+ participants had to self-report consistent SCD+ on any item from the ECog (again ≥3) as well as indicate worry/concern about their cognitive decline. For the final analysis, participants were considered SCD+ based only if they indicated worry/concern about their memory/thinking abilities, irrespective of their CCI or ECog scores.

### Structural MRI acquisition and processing

All participants were imaged using a 3T scanner with T1-weight imaging parameters (for more information see the ADNI’s official webpage for the MRI data acquisition protocol; http://adni.loni.usc.edu/methods/mri-tool/mri-analysis/). Baseline scans were download from the ADNI public website.

T1w scans for each participant were pre-processed through our standard pipeline in three steps: noise reduction (22), intensity inhomogeneity correction (23), and intensity normalization into range [0-100]. The pre-processed images were then both linearly (9 parameters: 3 translation, 3 rotation, and 3 scaling) (24) and nonlinearly (1 mm^3^ grid) (25) registered to the MNI-ICBM152-2009c average template (26). The quality of the linear and nonlinear registrations was visually verified by an experienced rater, and those that did not pass this quality control step were discarded (n = 7). The [^18^F]-AV45-PET data were downloaded from ADNI website.

### Deformation based morphometry

DBM analysis was performed to measure local anatomical differences in the brain by estimating the Jacobian determinant of the inverse nonlinear deformation field using MNI MINC tools. Jacobian determinant values reflect the volume of each voxel relative to the corresponding voxel on the average template. A DBM value of 1 indicates similar volume to the corresponding voxel in the template, and values lower (higher) than one indicates regions smaller (larger) than the same area in the template.

An atlas-based approach was also used to examine mean volume differences for ROIs (lateral ventricles, superior temporal, and amygdala from CerebrA atlas (26)) estimated by integrating the Jacobian of the deformation field within the ROI.

### SNIPE

Scoring by Nonlocal Image Patch Estimator (SNIPE) was used to measure the extent of Alzheimer’s disease related pathology using the linearly registered preprocessed T1-weighted images (27). SNIPE estimates a similarity metric for each voxel in the region of interest (e.g., hippocampus), indicating how much that voxel’s surrounding neighborhood resembles similar neighborhoods patches in a library of patients with probable Alzheimer’s disease or cognitively healthy controls. Average SNIPE values in the region of interest can then be used to examine the overall level of AD-related *macroscopic* neurodegeneration in that region. Positive SNIPE grading scores indicate normal appearing hippocampi, whereas negative scores indicate presence of AD-like atrophy.

### Statistical Analysis

All analyses were performed using MATLAB R2019b. Independent sample t-test were completed on the demographic information presented in Table 1. To investigate volume differences between SCD- and SCD+ participants we completed logistic regressions with ROI volume as the dependent variable and controlled for age, sex, and education. We performed separate logistic regression analyses for the four separate SCD definitions for each ROI. Additionally, we performed the analyses with and without *APOE ε4* and amyloid status.

Linear mixed effects models were conducted to examine whether SCD diagnosis would influence future longitudinal cognitive scores. A total of 1421 time points for 273 participants were included in the Alzheimer’s Disease Assessment Scale–Cognitive Subscale (ADAS-13) model and 820 timepoints for 273 participants were included in the Montreal Cognitive Assessment (MoCA) model:

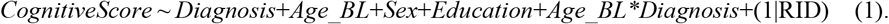

We explored the power of this model to examine the association between scores from the ADAS-13 and MoCA as the *CognitiveScore* and *Diagnosis*. The categorical variable of interest was Diagnosis, indicating SCD- or SCD+ status based on each definition. The models also included Time from baseline, sex, and years of education as covariates. Subject ID was included as a categorical random effect. All continuous variables were z-scored before being entered into the model. To express the unit change of ADAS and MoCA scores, the estimate from the model was then multiplied by the standard deviation of the scores divided by the standard deviation of the time at baseline factor.

Linear regressions were also completed to examine whether definition of SCD influenced amyloid levels between SCD- and SCD+. Five participants did not complete the PET imaging and therefore were not included in this analysis. A total of 270 participants were included in the model:

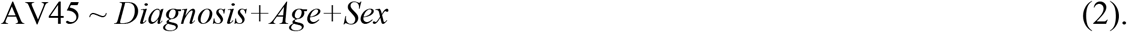

## Results

### Demographics

Table 1 shows the demographic information and cognitive testing scores for all participants. No statistically significant differences in mean age, education, or male:female ratio between groups was observed. Figure 1 displays the participant overlap between the different methods of defining SCD.

**Figure 1:**
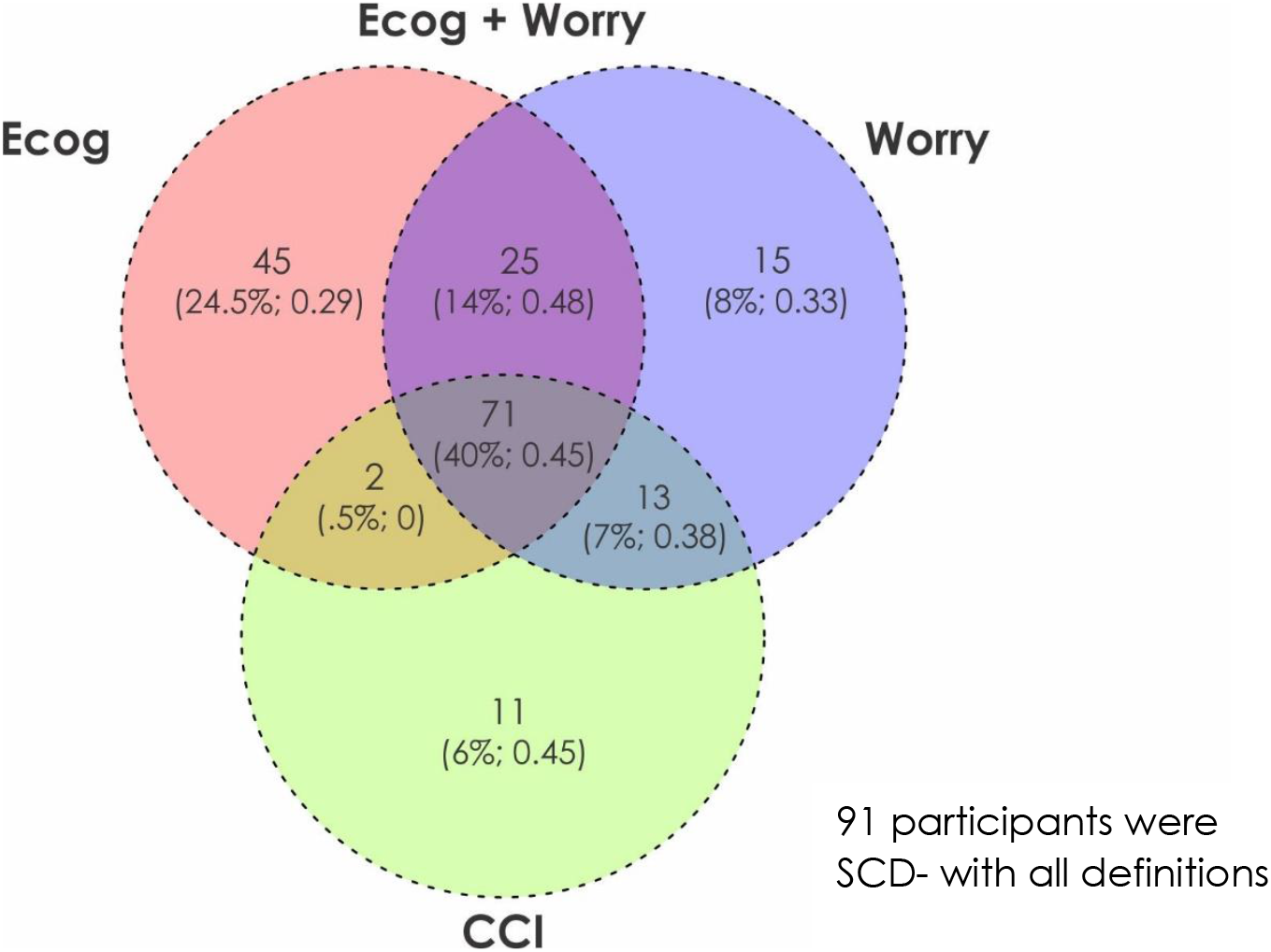
Venn Diagram representing the overlap of subjective cognitive decline (SCD) diagnosis between the four definitions of SCD. There was a total of 273 participants in the sample, 91/273 (33%) were SCD-with all definitions. The remaining 182 participants are shown in the Venn Diagram with the number of participants, the percentage of overall SCD+ sample, and the fraction of the sample that was amyloid positive. Overall, there were 72/182 (0.40) SCD participants that were amyloid positive. For the four SCD definitions, there were 97 SCD+ subjects defined by CCI, 143 SCD+ defined by ECog, 96 SCD+ defined by ECog &Worry and 124 defined by Worry only. Finally, only 40% of the SCD+ subjects are common between the four definitions.

### Atlas-based DBM analysis

Table 2 summarizes the results of the logistic regression models for both DBM and SNIPE analysis. Figure 2 shows significant t-statistic values obtained for the categorical diagnosis variable (Subjective Cognitive Decline; SCD+ and SCD-) for only the six regions tested (left/right hippocampus, amygdala, and superior temporal gyrus). Green regions indicate ROIs that were examined but were not significantly different between the groups. There were few anatomical differences detected by DBM. Only the right (OR = 0.72, 95% CI = -0.58 – -0.09, *p* = .008) and left (OR = 0.68, 95% CI = -0.64 – -0.01, *p* = .003) superior temporal regions were influenced by diagnosis when using the ECog-based definition. A trending effect of diagnosis was also observed in the right amygdala (OR = 0.79, 95% CI = -0.49 – 0.01, *p* = .064) for the ECog analysis. No other structures were significantly different for SCD+ and SCD-groups using the ECog definition. No structures were found to be different using the other SCD definitions. (All p-values reported after correction for multiple comparisons).

**Table 2:**
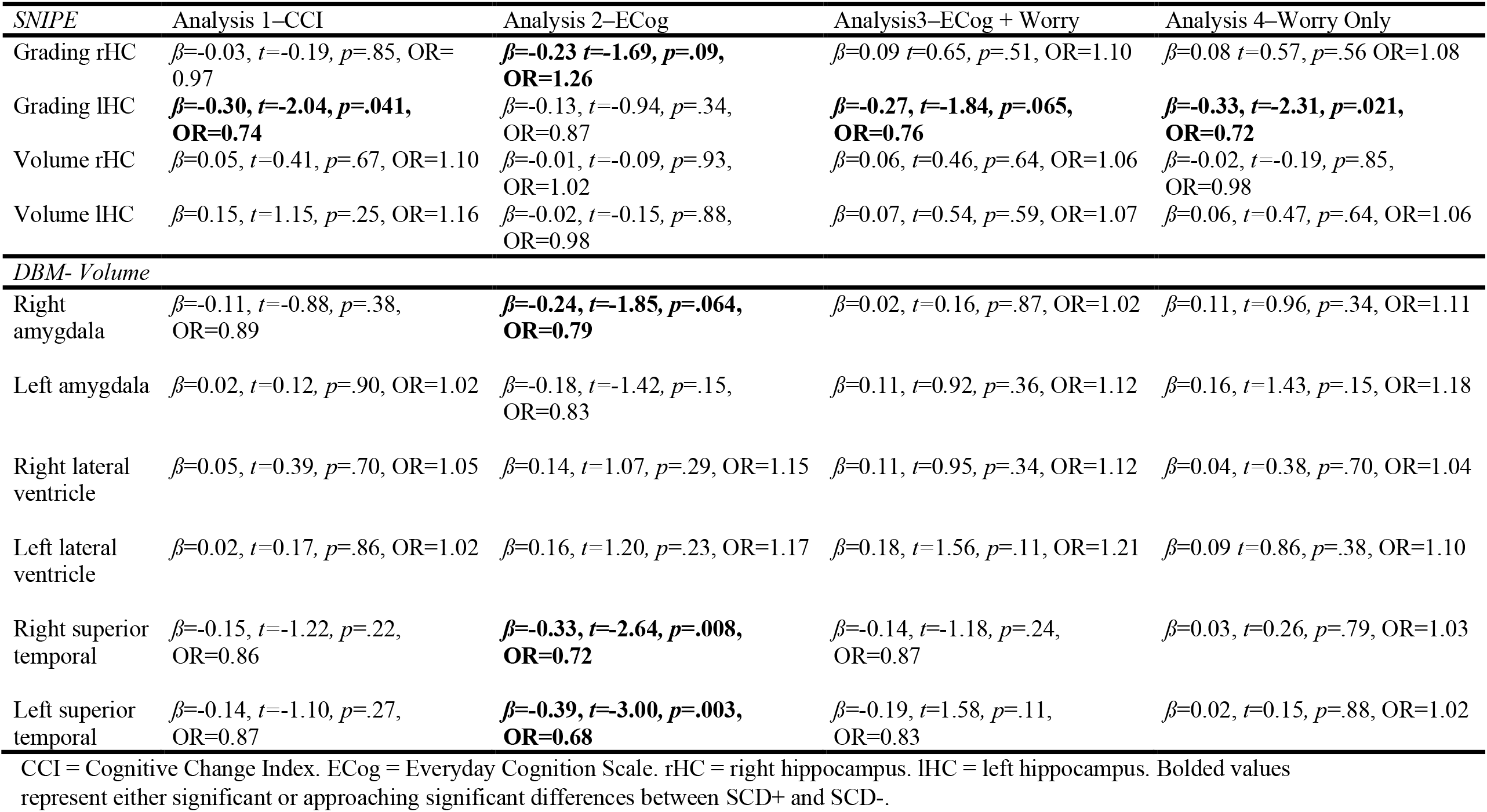
Logistic regression model results showing differences between cognitively healthy older adults with and without subjective cognitive decline

**Figure 2:**
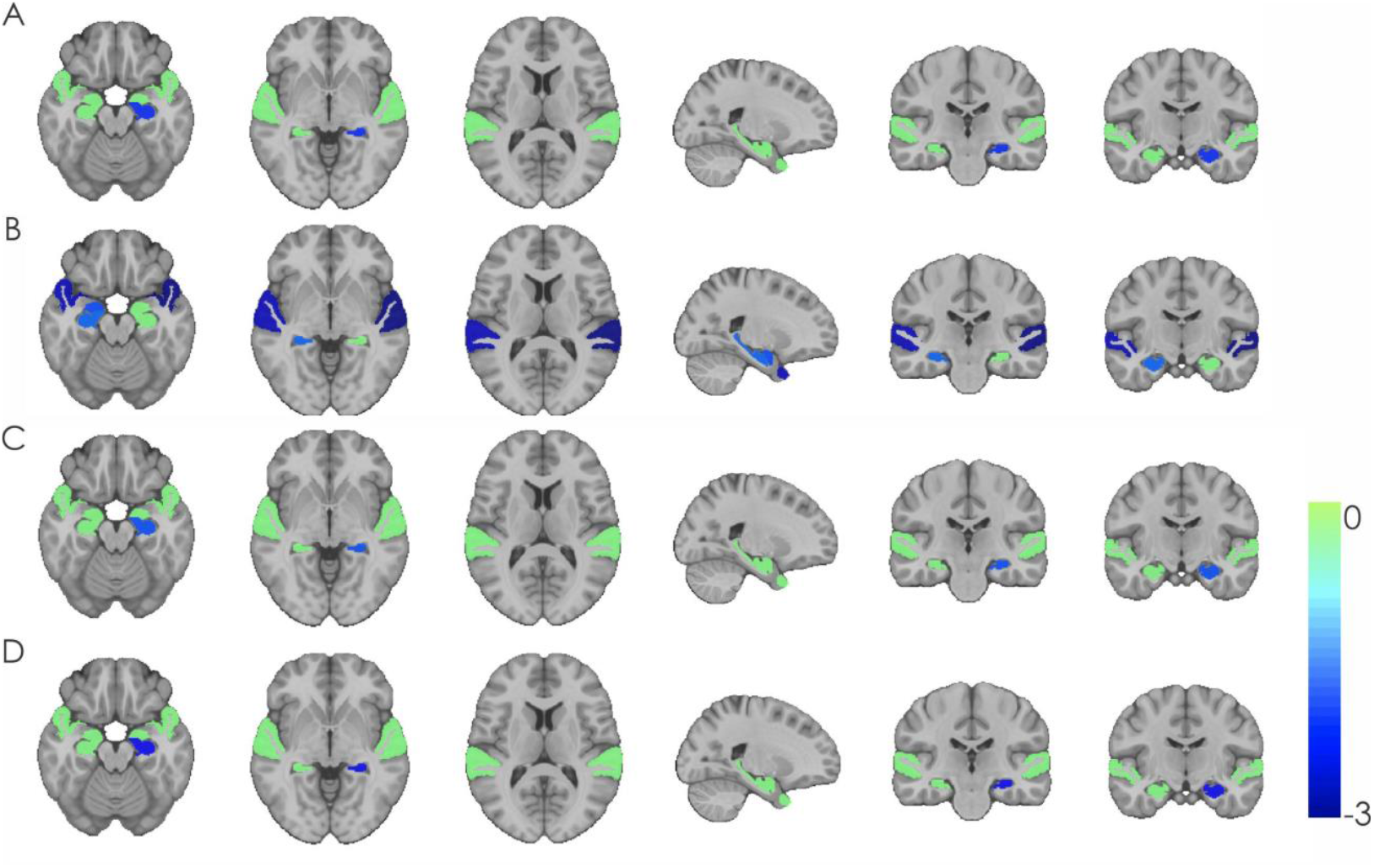
This figure shows significant t-statistic values obtained for the categorical diagnosis variable (Subjective Cognitive Decline; SCD+ and SCD-) for the six regions tested (left/right hippocampus, amygdala, and superior temporal gyrus). Green regions indicate ROIs that were examined but were not significantly different between the groups. Colder colors indicate lower DBM values in SCD+ compared to SCD-. A) Cognitive Change Index analysis – smaller left hippocampal grading in SCD+ vs SCD-. B) Everyday Cognition Scale analysis – smaller right hippocampal grading, right amygdala, and right and left superior temporal regions in SCD+ vs SCD-. C) Everyday Cognition Scale + Worry analysis – smaller left hippocampal grading in SCD+ vs SCD-. D) Worry analysis – smaller left hippocampal grading in SCD+ vs SCD-.

### SNIPE Analysis

There was an effect of diagnosis on grading in the left hippocampus for both the CCI definition of SCD (OR = 0.74, 95% CI = -0.60 – -0.01, *p* = .04) and Worry (OR = 0.72, 95% CI = -0.62 – - 0.05, *p* = .02), with an approaching significant effect for ECog + Worry definition (OR = -0.27, 95% CI = -0.56–0.02, *p* = .065).

### Amyloid & APOE Status

As can be observed in Table 3 when amyloid positivity and *APOE ε4* status were included in the models the differences due to diagnosis on volume did not significantly change. Amyloid positivity was significantly associated with volume for all SNIPE analyses, except for the ECog definition. On the other hand, *APOE* status was not associated with volume change in any of the ROIs or SCD definitions. As measured by AV-45, levels of amyloid did not differ between SCD- and SCD+ in any of the analysis (see Table 4). Levels of amyloid deposition also did not differ between the SCD+ groups using the four definitions. *APOE ε4+* status did not differ between SCD- and SCD+ populations or between SCD+ groups between the four definitions.

**Table 3:** Logistic regression model results including *APOE* and amyloid status showing the differences between cognitively healthy older adults with and without subjective cognitive decline

| *SNiPE* | Analysis 3–CCI | Analysis 2–ECog | Analysis3–ECog + Worry | Analysis 4–Worry Only |
| --- | --- | --- | --- | --- |
| Grading rHC | $\beta=-0.007, t=-0.05, p=.96, OR=0.99$<br>**Amyloid: $\beta=0.50, t=1.78, p=.075, OR=1.65$**<br>*APOE*: $\beta=-0.23, t=-0.76, p=.44, OR=0.80$ | **$\beta=0.24, t=1.76, p=.078, OR=1.28$**<br>Amyloid: $\beta=0.21, t=0.79, p=.43, OR=1.24$<br>*APOE*: $\beta=0.01, t=0.23, p=.98, OR=1.01$ | $\beta=0.12, t=0.81, p=.42, OR=1.13$<br>**Amyloid: $\beta=0.64, t=2.28, p=.022, OR=1.90$**<br>*APOE*: $\beta=-0.35, t=-1.14, p=.26, OR=0.70$ | $\beta=0.10, t=0.72, p=.47, OR=1.10$<br>**Amyloid: $\beta=0.50, t=1.87, p=.061, OR=1.66$**<br>*APOE*: $\beta=-0.16, t=-0.54, p=.59, OR=0.86$ |
| Grading lHC | **$\beta=-0.30, t=-1.98, p=.047, OR=0.74$**<br>Amyloid: $\beta=0.48, t=1.72, p=.085, OR=1.62$<br>*APOE*: $\beta=-0.23, t=-0.75, p=.45, OR=0.80$ | $\beta=-0.13, t=-0.92, p=.36, OR=0.88$<br>Amyloid: $\beta=0.17, t=0.63, p=.53, OR=1.18$<br>*APOE*: $\beta=-0.01, t=-0.05, p=.96, OR=0.99$ | **$\beta=-0.27, t=1.78, p=.075, OR=0.77$**<br>Amyloid: $\beta=0.61, t=2.16, p=.030, OR=1.83$<br>*APOE*: $\beta=-0.35, t=-1.15, p=.25, OR=0.70$ | **$\beta=-0.33, t=-2.26, p=.024, OR=0.72$**<br>Amyloid: $\beta=0.48, t=1.76, p=.078, OR=1.61$<br>*APOE*: $\beta=-0.17, t=-0.57, p=.57, OR=0.85$ |
| Volume rHC | $\beta=0.04, t=0.33, p=.74, OR=1.04$<br>**Amyloid: $\beta=0.50, t=1.78, p=.075, OR=1.64$**<br>*APOE*: $\beta=-0.22, t=-0.73, p=.46, OR=0.80$ | $\beta=-0.01, t=-.10, p=.91, OR=0.99$<br>Amyloid: $\beta=0.18, t=0.66, p=.50, OR=1.19$<br>*APOE*: $\beta=-0.01, t=-0.05, p=.96, OR=0.99$ | $\beta=0.05, t=0.36, p=.72, OR=1.05$<br>**Amyloid: $\beta=0.62, t=2.21, p=.027, OR=1.85$**<br>*APOE*: $\beta=-0.35, t=-1.13, p=.26, OR=0.71$ | $\beta=-0.03, t=-0.26, p=.79, OR=0.97$<br>**Amyloid: $\beta=0.49, t=1.83, p=.067, OR=1.63$**<br>*APOE*: $\beta=-0.17, t=-0.58, p=.56, OR=0.85$ |
| Volume lHC | $\beta=0.14, t=1.12, p=.26, OR=1.16$<br>**Amyloid: $\beta=0.49, t=1.77, p=.076, OR=1.64$**<br>*APOE*: $\beta=-0.22, t=-0.74, p=.46, OR=0.80$ | $\beta=-0.02, t=-.16, p=.87, OR=0.98$<br>Amyloid: $\beta=0.18, t=0.66, p=.51, OR=1.19$<br>*APOE*: $\beta=-0.13, t=-0.04, p=.96, OR=0.99$ | $\beta=0.06, t=0.49, p=.62, OR=1.07$<br>**Amyloid: $\beta=0.62, t=2.21, p=.027, OR=1.86$**<br>*APOE*: $\beta=-0.35, t=-1.14, p=.25, OR=0.70$ | $\beta=0.06, t=0.44, p=.66, OR=1.06$<br>**Amyloid: $\beta=0.49, t=1.82, p=.069, OR=1.63$**<br>*APOE*: $\beta=-0.16, t=-0.55, p=.58, OR=0.85$ |
| *DBM* | Analysis 3–CCI | Analysis 2–ECog | Analysis3–ECog + Worry | Analysis 4–Worry Only |
| Right amygdala | $\beta=-0.11, t=-0.82, p=.41, OR=0.90$<br>Amyloid: $\beta=0.41, t=1.48, p=.13, OR=1.51$<br>*APOE*: $\beta=-0.10, t=-0.35, p=.73, OR=0.90$ | **$\beta=-0.23, t=-1.83, p=.067, OR=0.79$**<br>Amyloid: $\beta=0.13, t=0.47, p=.64, OR=1.13$<br>*APOE*: $\beta=-0.02, t=-0.08, p=.94, OR=0.98$ | $\beta=0.02, t=0.18, p=.86, OR=1.02$<br>Amyloid: $\beta=0.13, t=0.52, p=.60, OR=1.14$<br>***APOE*: $\beta=-0.47, t=-1.72, p=.085, OR=0.62$** | $\beta=0.10, t=0.94, p=.35, OR=1.11$<br>Amyloid: $\beta=-0.01, t=-0.06, p=.95, OR=0.99$<br>*APOE*: $\beta=-0.31, t=-1.23, p=.21, OR=0.74$ |
| Left amygdala | $\beta=0.03, t=0.23, p=.82, OR=1.03$<br>Amyloid: $\beta=0.42, t=1.52, p=.13, OR=1.52$<br>APOE: $\beta=-0.10, t=-0.33, p=.74, OR=0.91$ | $\beta=-0.17, t=-1.39, p=.16, OR=0.84$<br>Amyloid: $\beta=0.12, t=0.45, p=.66, OR=1.13$<br>APOE: $\beta=-0.02, t=-0.06, p=.96, OR=0.98$ | $\beta=0.12, t=0.97, p=.33, OR=1.13$<br>Amyloid: $\beta=0.15, t=0.58, p=.56, OR=1.16$<br>**APOE: $\beta=-0.48, t=-1.75, p=.081, OR=0.62$** | $\beta=0.16, t=1.43, p=.15, OR=1.18$<br>Amyloid: $\beta=-0.01, t=-0.02, p=.98, OR=1.00$<br>APOE: $\beta=-0.31, t=-1.26, p=.21, OR=0.73$ |
| Right lateral ventricle | $\beta=0.04, t=0.30, p=.76, OR=1.04$<br>Amyloid: $\beta=0.41, t=1.48, p=.14, OR=1.51$<br>APOE: $\beta=-0.09, t=-0.30, p=.77, OR=0.91$ | $\beta=0.13, t=1.05, p=.29, OR=1.15$<br>Amyloid: $\beta=0.12, t=0.46, p=.64, OR=1.13$<br>APOE: $\beta=-0.02, t=-0.07, p=.95, OR=1.02$ | $\beta=0.10, t=0.80, p=.43, OR=1.10$<br>Amyloid: $\beta=0.12, t=0.49, p=.62, OR=1.13$<br>APOE: $\beta=-0.45, t=-1.65, p=.10, OR=0.63$ | $\beta=0.03, t=0.27, p=.79, OR=1.03$<br>Amyloid: $\beta=-0.03, t=-0.12, p=.90, OR=0.97$<br>APOE: $\beta=-0.30, t=-1.19, p=.23, OR=0.74$ |
| Left lateral ventricle | $\beta=0.01, t=0.10, p=.91, OR=1.01$<br>Amyloid: $\beta=0.42, t=1.50, p=.13, OR=1.51$<br>APOE: $\beta=-0.10, t=-0.32, p=.75, OR=0.91$ | $\beta=0.16, t=1.20, p=.23, OR=1.17$<br>Amyloid: $\beta=0.13, t=0.47, p=.64, OR=1.13$<br>APOE: $\beta=0.03, t=0.09, p=.93, OR=1.03$ | $\beta=0.17, t=1.39, p=.16, OR=1.18$<br>Amyloid: $\beta=0.12, t=0.48, p=.63, OR=1.13$<br>APOE: $\beta=-0.44, t=-1.58, p=.12, OR=0.65$ | $\beta=0.08, t=0.74, p=.46, OR=1.09$<br>Amyloid: $\beta=-0.03, t=-0.13, p=.89, OR=0.97$<br>APOE: $\beta=-0.28, t=-1.15, p=.25, OR=0.75$ |
| Right superior temporal | $\beta=-0.15, t=-1.21, p=.23, OR=0.86$<br>Amyloid: $\beta=0.41, t=1.50, p=.13, OR=1.51$<br>APOE: $\beta=-0.09, t=-0.30, p=.77, OR=0.92$ | **$\beta=-0.33, t=-2.64, p=.008, OR=0.72$**<br>Amyloid: $\beta=0.14, t=0.53, p=.60, OR=1.15$<br>APOE: $\beta=0.01, t=0.04, p=.97, OR=1.01$ | $\beta=-0.13, t=-1.19, p=.23, OR=0.87$<br>Amyloid: $\beta=0.11, t=0.44, p=.65, OR=1.21$<br>**APOE: $\beta=-0.47, t=-1.72, p=.085, OR=0.62$** | $\beta=0.02, t=0.23, p=.82, OR=1.03$<br>Amyloid: $\beta=-0.03, t=-0.11, p=.91, OR=0.97$<br>APOE: $\beta=-0.30, t=-1.21, p=.22, OR=0.74$ |
| Left superior temporal | $\beta=-0.14, t=-1.04, p=.30, OR=0.87$<br>Amyloid: $\beta=0.39, t=1.44, p=.15, OR=1.49$<br>APOE: $\beta=-0.06, t=-0.19, p=.85, OR=0.94$ | **$\beta=-0.39, t=-3.00, p=.002, OR=0.68$**<br>Amyloid: $\beta=0.10, t=0.36, p=.72, OR=1.10$<br>APOE: $\beta=0.11, t=0.37, p=.71, OR=1.11$ | $\beta=-0.18, t=-1.51, p=.13, OR=0.83$<br>Amyloid: $\beta=0.11, t=0.44, p=.65, OR=1.12$<br>**APOE: $\beta=-0.45, t=-1.66, p=.097, OR=0.63$** | $\beta=0.02, t=0.19, p=.85, OR=1.02$<br>Amyloid: $\beta=-0.03, t=-0.11, p=.91, OR=0.97$<br>APOE: $\beta=-0.30, t=-1.22, p=.22, OR=0.74$ |

CCI = Cognitive Change Index. ECog = Everyday Cognition Scale. rHC = right hippocampus. lHC = left hippocampus. Bolded values represent either significant or approaching significant differences between SCD+ and SCD-.

**Table 4:** Linear regression model results showing the group differences in AV-45 between SCD- and SCD+.

|  | Analysis 1–CCI | Analysis 2–ECog | Analysis3–ECog + Worry | Analysis 4–Worry Only |
| --- | --- | --- | --- | --- |
| Intercept | $\beta=0.71, SE= 0.13, t=5.41, p<.001$ | $\beta=0.71, SE= 0.13, t=5.56, p<.001$ | $\beta=0.71, SE= 0.13, t=5.43, p<.001$ | $\beta=0.71, SE= 0.13, t=5.41, p<.001$ |
| Group | $\beta=0.13, SE= 0.02, t=0.62, p=.54$ | $\beta=0.02, SE= 0.02, t=1.13, p=.26$ | $\beta=0.02, SE= 0.02, t=0.92, p=.35$ | $\beta=0.03, SE= 0.02, t=1.39, p=.17$ |
| Sex- Male | $\beta=-0.09, SE= 0.02, t=-4.14, p<.001$ | $\beta=-0.09, SE= 0.02, t=-4.14, p<.001$ | $\beta=-0.09, SE= 0.02, t=-4.15, p<.001$ | $\beta=-0.09, SE= 0.02, t=-4.08, p<.001$ |
| Age | $\beta=0.01, SE< 0.01, t=3.40, p<.001$ | $\beta=0.01, SE< 0.01, t=3.26, p<.001$ | $\beta=0.01, SE< 0.01, t=3.40, p<.001$ | $\beta=.01, SE< 0.01, t=3.38, p<.001$ |

CCI = Cognitive Change Index. ECog = Everyday Cognition Scale. Bolded values represent significant differences between SCD+ and SCD-.

### Cognitive Follow-up

Figure 3 displays the longitudinal change in cognitive scores for the ADAS and MoCA. For the ADAS, the only significant difference between SCD- and SCD+ was observed when using the CCI definition of SCD (ß=0.18, SE=0.09, *t=* 2.10, *p=*.036). This result indicates that people who were SCD+ had 0.40 units more of ADAS13 than those who were SCD. The model also revealed a significant effect of Age (ß=0.33, SE=0.04, *t=* 7.86, *p<*.001), Sex (ß=0.35, SE=0.09, *t=* 3.98, *p*<.001), and Education (ß=-0.09, SE=0.04, *t=* -2.23, *p=*.03). That is, with every year of increased age the ADAS13 increases by 0.73, there is a 0.77-unit difference between males and females, and every year of education increases ADAS13 scores by 0.20 units. The interaction between Time from Baseline and Diagnosis was not significant (*t*=1.31, *p*=.19). The other definitions of SCD were not associated with cognitive change as measured by the ADAS13.

**Figure 3.**
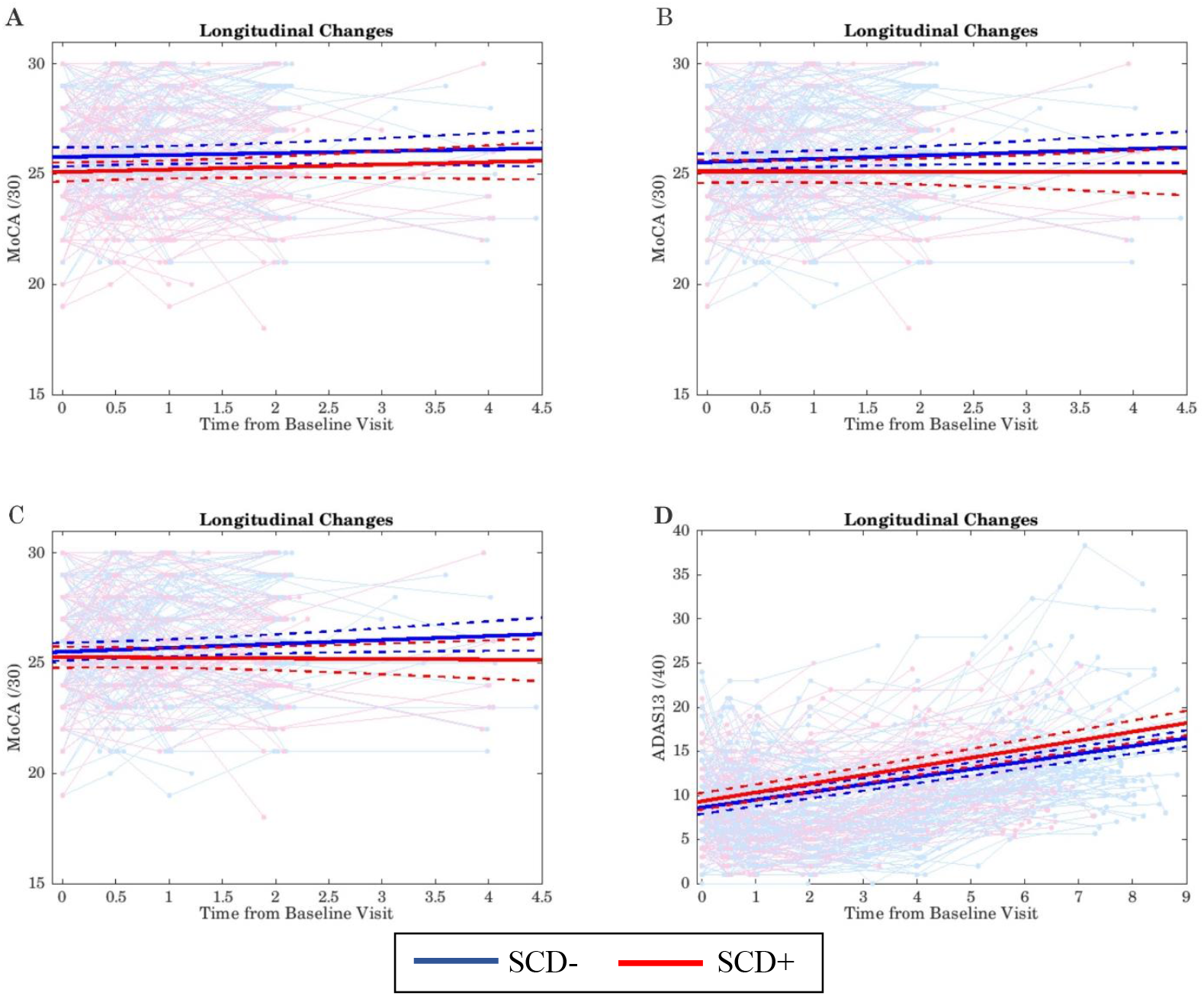
All figures show the longitudinal clinical change of each participant as well as the group change over time. Red lines = SCD+; Blue lines = SCD-. A) Longitudinal change of MoCA scores when defining SCD- and SCD+ based on ECog; B) Longitudinal change of MoCA scores when defining SCD- and SCD+ based on ECog + Worry; C) Longitudinal change of Montreal Cognitive Assessment (MoCA) scores when defining SCD- and SCD+ based on Worry; D) Longitudinal change of ADAS scores when defining SCD- and SCD+ based on the Cognitive Change Index

Longitudinal change in the MoCA was observed for the ECog definition of SCD (ß= -0.27, SE=0.09, *t=* 2.87, *p=*.004). People who were SCD+ had 0.77 units less of MoCA than those who are SCD-. This model also revealed a significant effect of Age (ß=-0.28, SE=0.05, *t=* -6.03, *p<*.001), Sex (ß=-0.31, SE=0.10, *t= -*3.323, *p*=.001), and Education (ß=0.14, SE=0.05, *t=* 2.87, *p=*.004). The interaction between Time from Baseline and Diagnosis was not significant (*t*<1, *p*=.87). Every year of increased age reduces the MoCA score by 0.80 units, there is a difference of 0.88 units between males and females, and every year of education increases MoCA scores by 0.40 units.

The ECog + Worry definition of SCD also revealed a significant group effect (ß= -0.22, SE=0.10, *t=* -2.18, *p=*.03). That is, people who were SCD+ had 0.63 units less of MoCA than those who are SCD-. Similar to the ECog results, this model also revealed a significant effect of Age (ß=-0.29, SE=0.05, *t=* -5.94, *p<*.001), Sex (ß=-0.34, SE=0.10, *t= -*3.39, *p<*.001), and Education (ß=0.14, SE=0.05, *t=* 2.90, *p*=.004). The interaction between Time from Baseline and Diagnosis was not significant (*t*=-1.10, *p*=.27). Every year of increased age reduces the MoCA score by 0.83 units, there is a difference of 0.97 units between males and females, and every year of education increases MoCA scores by 0.40 units. The CCI and Worry definition of SCD was not significantly associated with MoCA scores.

## Discussion

The SCD Initiative working group has suggested standards for defining SCD in research (6). Nevertheless, current studies have used widely discrepant methodologies for defining SCD, making generalizations difficult. This lack of standardization may contribute to some of the inconsistencies in the current literature examining if people with SCD exhibit AD-related pathology. The current study was designed to examine these inconsistencies by comparing four methods of defining SCD using novel MRI analysis techniques. The current study investigated three main questions: 1) What is the overlap of SCD+ participants defined using the four different SCD definitions? 2) do SCD- and SCD+ atrophy differences vary depending on the definition of SCD? and 3) are longitudinal cognitive trajectories of SCD- and SCD+ populations different between the four definitions of SCD? We observed that the four methods do not categorize older adults similarly as there was little overlap between the definitions. Both atrophy differences and longitudinal cognitive trajectories between SCD- and SCD+ vary depending on the method used to define SCD.

First, only 40% of the SCD+ participants were common between the four definitions. This result suggests that there are major inconsistencies in who is identified as having SCD between these methods. Such differences in identifying who has SCD can be a problem if a clinician is attempting to determine which measure to use to predict/screen for AD or future cognitive decline. With little overlap between these methods, clinicians may screen out people who could have preclinical AD or may include people who do not have preclinical AD. Therefore, if early treatment becomes available, they could miss treatment for some patients while providing treatment that is not necessary to others. Relying on subjective clinical judgment may introduce further inconsistencies across care providers and clinics. To better understand which SCD definition best predicts future brain atrophy and cognitive decline more research is needed to standardize how SCD is screened for, measured, and diagnosed.

Second, we observed SCD-:SCD+ volume differences between groups using the four definitions. However, these group effects were not in the same locations across the definitions. Hippocampal volume was smaller in SCD+ relative to SCD-when using both the CCI and Worry and approached significance for the ECog + Worry definition. Decreased hippocampal volumes in those with SCD+ compared to SCD-in these two methods are consistent with other studies (11,28–30). However, we did not find hippocampal volume group differences when defining SCD with the ECog. This findings is *also* consistent with several studies that did not report hippocampal atrophy in people with SCD (12,14,31). These differences provide insight into how different definitions of SCD may influence atrophy differences observed between SCD+ and SCD-.

The ECog was sensitive to SCD-:SCD+ group differences in the right and left superior temporal regions that were not observed with the other definitions. Previous studies have found that white matter (20) and cortical thickness (32) in the superior temporal regions are sensitive to progression to amnestic MCI and AD. Yue et al. observed that enlarged white matter at the banks of the superior temporal sulcus was associated with increased progression from SCD to MCI over 7 years (20). Eskildsen et al. found that cortical thickness of the left superior temporal sulcus was a key feature in discriminating between people with MCI who remain stable and people with MCI who convert to AD over 3 years (32). It is thus possible that the ECog may be sensitive to early decline several years in advance.

Third, we found different cognitive trajectories between the SCD+ groups using the four definitions. While an association between the CCI and future cognitive decline as measured by the ADAS-13 was found, the CCI was not associated with cognitive change on the MoCA. On the other hand, the ECog definition of SCD was associated with decline on the MoCA, but not on the ADAS-13. It is possible that these differences are associated with the type of cognitive decline the questionnaires are sensitive to. For both the ADAS and MoCA models, we found that higher education was associated with better cognitive scores. We also found that males and older age was associated with worse performance. The worry definition was not associated with future cognitive decline as measured by either the ADAS or MoCA. This finding is further supported by a less significant effect observed for the association between ECog + Worry and MoCA scores than for the association between ECog alone and MoCA; suggesting that the ECog definition is driving the association between SCD and cognitive change for MoCA scores.

The mixed group volume differences and cognitive trajectories between SCD- and SCD+ observed between the four definitions of SCD suggest these methods may measure different *construct*s or types of SCD. This hypothesis is further supported by the participant overlap between the methods reaching only 40% and the different patterns of atrophy in the SCD+ groups. The CCI and Worry methods revealed hippocampal volume declines in people with SCD. These two methods may tap into subjective cognitive declines involved with the left hippocampus such as verbal memory deficits (33). Hippocampal declines are observed early in AD disease progression. Therefore, the CCI and Worry methods may be sensitive to SCD related to AD. When also taking into consideration the cognitive trajectories of CCI and Worry, only the CCI was associated with future cognitive decline. These findings suggest that the CCI questionnaire may be the most sensitive (of the four measures employed in this study) to preclinical AD. Reduced volume in the superior temporal region was observed in SCD+ relative to SCD- but only using the ECog definition. Thus, the ECog may be sensitive to cognition declines related to the superior temporal gyrus such as episodic memory coding, language comprehension, and speech processing (34,35). Although declines in the superior temporal region have been observed as a sensitive factor for future cognitive decline, this region is not a key component underlying AD-related atrophy. It is possible that the ECog is sensitive to cognitive decline (as measured by the MoCA) associated with another dementia other than AD. Taken together, the ECog and CCI may be sensitive to different subtypes of SCD. Another recent study also identified multiple SCD subgroups, each characterized by unique patterns of brain atrophy (36).

It is important to note that *all* the methods of defining SCD in this sample had a similar ratio of amyloid positivity, *APOE ε4+*, and tau levels in the participants. According to the recent National Institute on Aging Alzheimer’s Association (NIA-AA), SCD is part of the transitional “Stage 2” of AD. In this stage people should exhibit pathological tau and Aß markers. In the current study *none* of methods used to define SCD resulted in SCD+ groups with higher levels of pathology than the SCD-groups. This finding suggests that although the CCI appears to be sensitive to AD-related atrophy and cognitive decline, it may not be correctly identifying those with tau and amyloid pathology.

It should be noted that the image processing employed in this study (i.e., DBM and SNIPE) have been developed and extensively validated for use in multi-center and multi-scanner studies. These processing methods have previously shown patterns of atrophy in cognitively normal, MCI, dementia, and neurodegenerative disease populations, including ADNI (37–40). These techniques thus have the required sensitivity to reveal group differences between SCD- and SCD+. Therefore, the lack of SCD-:SCD+ group differences observed in this study is not the result of image processing methods that are not sensitive to observe group differences.

There are a few limitations of the current study. First, in current study we used cross-sectional MRI data. Future research should use a longitudinal design to determine if the conversion rate to MCI from SCD varies in the assigned groups with all four definitions. A longitudinal study would not only help differentiate between the subtypes of SCD but would also improve our understanding of how the questionnaires may be associated with regional atrophy in SCD+ vs. SCD-. In the current dataset, the CCI was only administered at screening and thus a follow-up using a longitudinal design with the CCI was not possible. Another limitation of the current study is the use of a population-based cohort. This study design is a limitation because people who seek medical help (i.e., memory clinic consultation) for memory concerns show more hippocampal atrophy than those who do not seek help (41) and may be more likely to convert to MCI (17). Within our sample, all our participants education levels were quite high. High education is a limitation because the changes observed in this sample may not be representative of all populations.

## Conclusion

The current study was the first to compare four commonly used methods of defining SCD in the same sample of participants. In addition, we employed a novel MRI analysis technique which has been found to be sensitive to AD-related cognitive decline. This study observed that regional atrophy and future cognitive decline observed in SCD+ populations will depend on the SCD definition used. Although hippocampal and superior temporal volumes differed between SCD- and SCD+, these differences were dependent on the SCD definition employed. Furthermore, cognitive trajectories also varied between the SCD definitions. The CCI appears to better identify SCD+ people with early cognitive decline related that may be related to AD-pathology. This result is further supposed by the SCD+ group identified by the CCI also exhibiting smaller left hippocampal grading than SCD-. The ECog questionnaire was also associated with future cognitive decline, but with atrophy in the superior temporal regions as opposed to the hippocamps, suggesting that the ECog may be sensitive to another type of cognitive decline.

These findings have significant implications for both clinicians and researchers. In both clinical and research settings, it is crucial to use a definition which has high sensitivity and specificity to identify individuals in the preclinical stages of AD. Misidentifying people who will progress to AD reduces the likelihood of researchers finding an early biomarker and reduces the chances of clinicians providing effective treatments to slow or stop AD progression. Future research needs to determine which questionnaire is sensitive to atrophy, future cognitive decline, and tau and amyloid pathology to be successful at accurately predicting who will progress to AD.

## Data Availability

Data used in preparation of this article were obtained from the Alzheimer's Disease Neuroimaging Initiative (ADNI) database (adni.loni.usc.edu).

## Acknowledgments

Data collection and sharing for this project was funded by the Alzheimer’s Disease Neuroimaging Initiative (ADNI) (National Institutes of Health Grant U01 AG024904) and DOD ADNI (Department of Defense award number W81XWH-12-2-0012). ADNI is funded by the National Institute on Aging, the National Institute of Biomedical Imaging and Bioengineering, and through generous contributions from the following: AbbVie, Alzheimer’s Association; Alzheimer’s Drug Discovery Foundation; Araclon Biotech; BioClinica, Inc.; Biogen; Bristol-Myers Squibb Company; CereSpir, Inc.; Cogstate; Eisai Inc.; Elan Pharmaceuticals, Inc.; Eli Lilly and Company; EuroImmun; F. Hoffmann-La Roche Ltd and its affiliated company Genentech, Inc.; Fujirebio; GE Healthcare; IXICO Ltd.; Janssen Alzheimer Immunotherapy Research & Development, LLC.; Johnson & Johnson Pharmaceutical Research & Development LLC.; Lumosity; Lundbeck; Merck & Co., Inc.; Meso Scale Diagnostics, LLC.; NeuroRx Research; Neurotrack Technologies; Novartis Pharmaceuticals Corporation; Pfizer Inc.; Piramal Imaging; Servier; Takeda Pharmaceutical Company; and Transition Therapeutics. The Canadian Institutes of Health Research is providing funds to support ADNI clinical sites in Canada. Private sector contributions are facilitated by the Foundation for the National Institutes of Health (www.fnih.org). The grantee organization is the Northern California Institute for Research and Education, and the study is coordinated by the Alzheimer’s Therapeutic Research Institute at the University of Southern California. ADNI data are disseminated by the Laboratory for Neuro Imaging at the University of Southern California.

MD is supported by a scholarship from the Canadian Consortium on Neurodegeneration in Aging as well as an Alzheimer Society Research Program (ASRP) postdoctoral award. The Consortium is supported by a grant from the Canadian 24 Institutes of Health Research with funding from several partners including the Alzheimer Society of Canada, Sanofi, and Women’s Brain Health Initiative.

## Footnotes

1 In this manuscript SCD+ refers to cognitive normal older adults with subjective cognitive decline and is not referring to SCD *plus* as defined by Jessen et al. (6).

